# Explainable AI enables clinical trial patient selection to retrospectively improve treatment effects in schizophrenia

**DOI:** 10.1101/2021.01.11.20248788

**Authors:** Monika S. Mellem, Matt Kollada, Jane Tiller, Thomas Lauritzen

## Abstract

**Background:** Heterogeneity among patients’ responses to treatment is prevalent in psychiatric disorders. Personalized medicine approaches – which involve parsing patients into subgroups better indicated for a particular treatment – could therefore improve patient outcomes and serve as a powerful tool in patient selection within clinical trials. Machine learning approaches can identify patient subgroups but are often not “explainable” due to the use of complex algorithms that do not mirror clinicians’ natural decision-making processes.

**Methods:** Here we combine two analytical approaches – Personalized Advantage Index and Bayesian Rule Lists – to identify paliperidone-indicated schizophrenia patients in a way that emphasizes model explainability. We apply these approaches retrospectively to randomized, placebo-controlled clinical trial data to identify a paliperidone-indicated subgroup of schizophrenia patients who demonstrate a larger treatment effect (outcome on treatment superior than on placebo) than that of the full randomized sample as assessed with Cohen’s d. For this study, the outcome corresponded to a reduction in the Positive and Negative Syndrome Scale (PANSS) total score which measures positive (e.g., hallucinations, delusions), negative (e.g., blunted affect, emotional withdrawal), and general psychopathological (e.g., disturbance of volition, uncooperativeness) symptoms in schizophrenia.

**Results:** Using our combined explainable AI approach to identify a subgroup more responsive to paliperidone than placebo, the treatment effect increased significantly over that of the full sample (p<0.0001 for a one-sample t-test comparing the full sample Cohen’s d=0.82 and a generated distribution of subgroup Cohen’s d’s with mean d=1.22, std d=0.09). In addition, our modeling approach produces simple logical statements (*if-then-else*), termed a “rule list”, to ease interpretability for clinicians. A majority of the rule lists generated from cross-validation found two general psychopathology symptoms, disturbance of volition and uncooperativeness, to predict membership in the paliperidone-indicated subgroup.

**Conclusions:** These results help to technically validate our explainable AI approach to patient selection for a clinical trial by identifying a subgroup with an improved treatment effect. With these data, the explainable rule lists also suggest that paliperidone may provide an improved therapeutic benefit for the treatment of schizophrenia patients with either of the symptoms of high disturbance of volition or high uncooperativeness.

**Trial Registration:** clincialtrials.gov identifier: NCT 00083668; registered May 28, 2004

## 1. Background

The primary goal in a placebo-controlled clinical trial testing the efficacy of an experimental medication is to show a treatment effect – that patients randomized to receive the medication have improved outcomes compared to those receiving placebo. Within psychiatry, there is heterogeneity in patients’ responses, however, with some not responding well or at all [e.g., 1; 2] which can weaken the overall response of the treatment-receiving group compared to placebo. Additionally, the placebo response is robust in psychiatric disorders [3] making assessments of treatment efficacy more difficult. A method termed Personalized Advantage Index (PAI) has been recently developed to uncover subgroups of patients, termed “treatment-indicated,” may be more responsive to a particular treatment than placebo and that predictive modeling could lead to personalized medicine approaches for subtyping treatment-indicated patients [4]. In particular, this could also help improve patient selection for clinical trials of that medication to enrich for patients most likely to show a treatment effect.

One of the limitations in using PAI to improve patient selection for clinical trials is insufficient explainability in how the model makes its decisions, as explainability is a critical attribute for a clinician to consider using an algorithm for patient selection. Prior work in depression has used the PAI approach to identify the most predictive variables [5], but interpretability of the models for clinicians could be further improved as they would require interpretation of regression coefficients and do not suggest clear cutoffs for predictor variables. Here, we additionally used an approach inspired by explainable artificial intelligence (XAI), the Bayesian Rule Lists algorithm (BRL) [6, 7], to both help identify the most predictive variables and explain those predictions of treatment-indicated patients from PAI with simple *if-then-else* statements that better mirror a clinician’s decision-making process by using Boolean criteria with clear cutoffs for predictor variables. The combined analytical approach of PAI and BRL was previously tested in depression and found to retrospectively identify a subgroup with improved treatment effect for the novel antagonist BTRX-246040 [8]. But it has yet to be tested in other psychiatric populations.

While improving patient selection of a clinical trial is one potential use, it is important to note the constraints on this PAI and BRL approach for this goal and some additional opportunities. This approach requires both baseline and post-treatment (or imputed post-treatment since patients often do not discontinue at random) measurements of patients receiving a particular treatment in order to learn the baseline characteristics of a treatment-indicated subgroup for this treatment prior to enrollment, so it may not be appropriate for clinical trial patient selection when no similar trial has been performed. One opportunity is thus to use it to learn the optimal subgroup from a negative clinical trial (as in [8]) and re-launch a more targeted clinical trial of the same treatment using the algorithm to identify a treatment-indicated subgroup. Enrolling this subgroup could help increase the treatment effect size of the more targeted clinical trial. A second opportunity is in using this approach to develop a decision-making support system for clinicians to prescribe medications already on the market to a targeted subgroup likely to have increased treatment efficacy. The data requirements of the PAI and BRL approach could be satisfied in all these scenarios.

In this study, we present this combined PAI and BRL approach to patient selection for clinical trials using XAI and validation in schizophrenia patients through a retrospective analysis of a clinical trial. By showing that baseline features alone can be used to identify a subgroup of patients who demonstrate a larger average treatment effect, this approach opens up the possibility of identifying a targeted subgroup of patient prior to randomization to an arm and improving the clinical trial outcome by using this patient subgroup in particular.

## 2. Methods

### 2.1 Study

We analyzed data from a 6-week randomized, double-blind, placebo-controlled study evaluating the efficacy of extended-release paliperidone in the treatment of patients with schizophrenia (clincialtrials.gov identifier: NCT 00083668). This trial was a success, and paliperidone currently has FDA approval in this population. This study makes use of de-identified data made available via the YODA Project (https://yoda.yale.edu/; research proposal number 2019-4080) and was exempt from ethical oversight.

Patients were assessed for eligibility and phenotyped using standard clinical assessments at baseline and randomized to one of several arms. In this analysis, we used data from the 15mg/day paliperidone arm (n=113) and the placebo arm (n=120). We selected the 15mg/day arm over arms testing lower doses (3mg or 9mg per day) as it gave the greatest efficacy effect compared to placebo. The efficacy endpoint was the Positive and Negative Syndrome Scale (PANSS), a set of 30 questions administered by a trained clinician and scored on a 1-7 ordinal scale (7 is most severe). The PANSS was administered weekly. After dropping patients with missing baseline values used as features in the modeling (see below), 95 patients remained in the treatment arm and 102 in the placebo arm.

### 2.2 Modeling

Our approach to identify treatment-indicated patients and improve the explainability of the machine learning algorithm output classifying these patients involved combining two approaches. First, we used the Personalized Advantage Index (PAI) algorithm [4] to create an index/score that is then used to label a patient as treatment-indicated or rest-indicated (the rest of the subjects who are not treatment-indicated). This machine learning approach relies on multiple linear regression which provides some level of explainability through the coefficients of the predictors but is likely more complex than clinicians’ decision making processes which are closer in form to decision trees or lists. Thus, in a second step, we used the Bayesian Rule Lists classifier [6, 7] along with the predicted treatment- and rest-indicated labels from PAI to create a more explainable classifier using decision lists. The overview of the workflow is presented in Figure 1.

**Figure 1:**
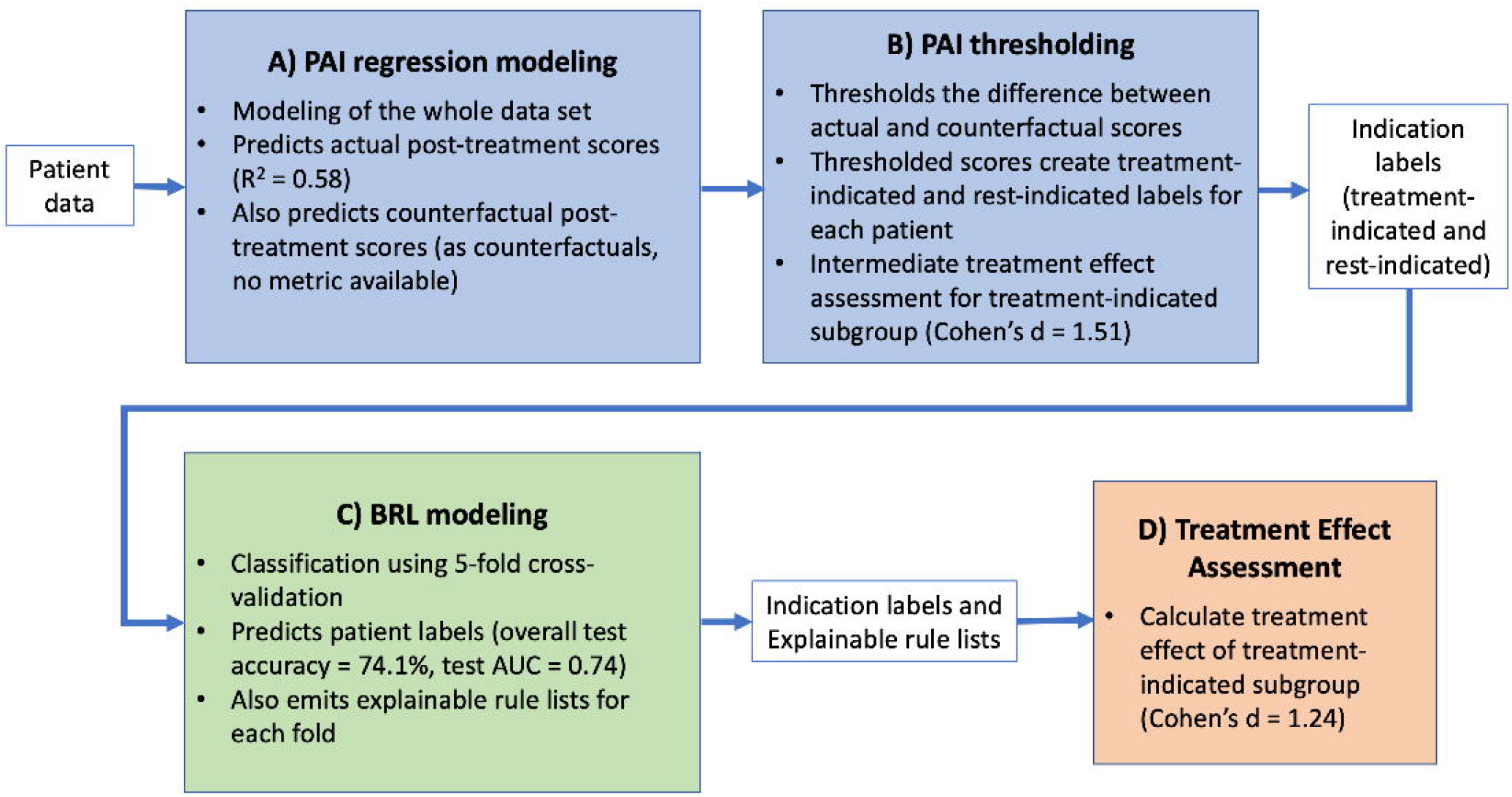
Overview of PAI and BRL modeling workflow. **A)** The first step is PAI regression modeling which takes in data listed in Table 1 and trains on the whole data set to predict both actual and counterfactual post-treatment scores for individual patients (actual scores can be compared with predicted actual scores and resulted in an R^2^ = 0.58 as shown in Figure 2). **B)** The PAI Thresholding step thresholds the difference between actual and counterfactuals to create indication labels for each patient. A treatment-indicated subgroup had a treatment effect size of Cohen’s d= 1.51 as an intermediate assessment, but explainability of model decisions needs improvement, so the BRL step addresses this need. **C)** The BRL modeling uses 5-fold cross-validation to assess generalization ability to unseen samples. The predictions generated for test samples over all folds had an accuracy of 74.1% and an AUC of 0.74 for this classifier. Importantly, it emits an explainable rule list for each fold. **D)** The final step is assessing the treatment effect of the treatment-indicated subgroup identified by BRL (Cohen’s d = 1.24 as seen in Figure 5).

#### 2.2.1 Personalized Advantage Index Modeling and Labeling

In our modeling approach, the PAI algorithm was first used to identify treatment-indicated patients. The PAI approach predicts actual and counterfactual outcomes (i.e., a patient’s outcome for their assigned arm, drug or placebo, and the non-assigned arm) and calculates the difference between these two scores (as previously described in [4, 5]).

**Table 1:** Model inputs and outputs. Several sequential outputs are generated during PAI modeling and are listed in order of generation.

| Model Input Features | PAI Model Output | BRL Model Output |
| --- | --- | --- |
| <u>Demographics</u><br>• Age<br>• Sex<br><u>Symptom scales</u><br>• 30 Baseline individual item PANSS scores<br>• Baseline total PANSS score<br>• Baseline daytime drowsiness and quality of sleep scores from the Sleep Visual Analog Scale (VAS)<br>• Baseline Personal and | • Predicted actual and counterfactual week 6 PANSS scores<br>• Numerical PAI score (predicted outcome on treatment – predicted outcome on placebo)<br>• Labels: treatment-indicated, rest-indicated created from thresholded PAI score | • Labels: treatment-indicated and rest-indicated |
| Social Performance Scale score<br>• Baseline total Schizophrenia Quality of Life Scale score<br><u>Others</u><br>• Randomized arm (treatment or placebo; PAI only)<br>• Interactions of randomized arm with the other features (PAI only). |  |  |

Briefly, we used an Elastic Net regressor (implemented in the python package scikit-learn), with a grid search for hyperparameter optimization across the range of *alpha* = [0.001, 0.01, 0.1, 1, 10] and *l1*_*ratio* = [0.1, 0.5, 0.9]. The input features are listed in Table 1 where “baseline” refers to week 0 of the trial, which precedes treatment arm randomization. The outcome modeled was the 6-week post-treatment total PANSS score. Scores from patients missing this 6-week score were replaced with their last observation (n = 80 patients) which was consistent with the approach used in the original clinical trial analysis. This multiple regression model then predicts the actual post-treatment PANSS score (on the patient’s randomized arm) and the hypothetical counterfactual score (by substituting the other arm in the regression equation). PAI then returns a quantitative score for each patient that indicates the difference between these predicted drug and placebo outcomes with better performance on drug corresponding to a negative PAI score. A subsequent threshold then creates the two classes of treatment-indicated and rest-indicated with possible thresholds examined in descending steps of 0.5 (0, −0.5, −1, −1.5,…). We selected the threshold that allocates ∼50% of the sample to treatment-indicated class to maintain a balanced data set for the BRL classifier. Please note that selecting a PAI score threshold is a matter of balancing algorithmic needs (two-class classifiers work best with balanced data), and clinical needs (clinicians may consider a specific percent decrease in symptoms, such as at least 30%, a clinically-meaningful decrease), and researchers who use this method in the future may need to reassess whether the 50% criteria used here will work in their scenario. As PAI was responsible for generating the best possible indication labels for BRL to train on (i.e., generating “ground truth” labels for BRL), the PAI regression model was trained on all the data. Please note that these are relative ground-truth labels – the best labels we can come up with but not perfect since the real ground-truth of counterfactual predictions will never be known. Thus these labels function as ground-truth for training BRL, as true ground-truth labels cannot be known with this clinical trial design. The grid search of hyperparameters showed that *alpha* = 0.1, *l1*_*ratio* = 0.1 minimized the R^2^.

#### 2.2.2 BRL Modeling and Labeling for Additional Explainability

The BRL algorithm was used to create a more explainable model from the initial PAI treatment-indicated and rest-indicated results. BRL uses sequenced logical rules and Bayesian inference to make classifications [6, 7]. Here, we took in the same baseline features other than the randomized arm and interactions (Table 1) and classified patients as treatment- or rest-indicated using the BRL-generated *if-then-else* statements. A Bayesian rule list is composed of Boolean statements that evaluate if features fit certain criteria such as “If depression symptom score > 10” and the subsequent classification if the statement is true – “then, patient is treatment-indicated.” These statements are closer to a physician’s decision-making process than are the PAI regression outputs (feature coefficients without clear cutoffs for feature values). Hyperparameters were set at 3 for the max rule length (number of Boolean statements combined in an individual rule), 2 for the Bayesian prior hyperparameters of expected individual rule length and 2 for the expected rule list length (excluding the final base case). We used a 5-fold cross-validation framework that generated a model (a rules list) on the 80% of training data and used it to classify the patients in the remaining 20% of test data as treatment- or rest-indicated. Thus five rules lists were generated from the five folds of cross-validation. As an additional output of this study, we generated a “final” BRL model by training the model on the full data set. While we have not tested this final BRL patient selection tool here on an external data set, others could use it if they have the appropriate data.

#### 2.2.3 Comparing Treatment Effects

After classifying each patient as treatment- and rest-indicated labels using BRL, we assessed if treatment-indicated patients showed an improved treatment effect compared with the full sample. It is important to note that as the treatment-indicated patients were randomized to both drug and placebo arms, we were able to evaluate their actual post-treatment outcomes and to calculate their group-level treatment effect. Here, the treatment effect was assessed on the actual week 6 PANSS total scores of individual patients grouped by their treatment arm (using Cohen’s d as a measure of treatment effect size). After determining the labels from the BRL outputs, the actual week 6 PANSS total scores for the treatment-indicated patient subgroup were used to calculate the treatment effect for that subgroup. Then the treatment effect of the treatment-indicated subgroup was compared with the treatment effect of the full randomized sample. Our null hypothesis is that patient selection with BRL provides no improvement of the treatment effect, while our alternative hypothesis is that BRL does improve the treatment effect relative to that of the full sample. Thus to test this statistically, we generated a distribution treatment effects by performing BRL 100 times and calculating the Cohen’s d for each BRL-labeled treatment-indicated subgroup. We then compared this distribution of 100 Cohen’s d’s with the Cohen’s d of the full sample using a two-sided, 1-sample t-test. Additionally, we assessed the consistency of classification for each subject across the five rule lists generated from the five folds of cross-validation.

## 3. Results

Our approach to identify treatment-indicated patients and improve the explainability of the machine learning algorithm output classifying these patients involved combining two approaches – PAI and BRL. While the final output and results are the treatment-labeled patient subgroup and the *if-then-else* rules list from the BRL model, the initial modeling with the PAI algorithm produced some interim results that we first examined. The PAI regression equation modeled actual week 6 PANSS scores using actual treatment arm assignment and several demographic and baseline symptom severity predictor variables (see Table 1). Figure 2 shows the comparison of measured week 6 PANSS scores with predicted scores (a perfect model would show all samples sitting on the x=y line). The R^2^ of the model shows it explained 58% of the variance (adjusted R^2^ = 0.32). Then, by substituting in the counterfactual randomization arm, the regression equation was used to make predictions of the week 6 PANSS scores if patients were receiving the counterfactual treatment. The difference between actual and counterfactual predictions were used to calculate the PAI scores (predicted score on treatment – predicted score on placebo) and determine the indication labels to be used by BRL. The distribution of PAI scores are shown in Figure 3, and a threshold of −9.5 generated two balanced classes: treatment-indicated (n=100) and rest-indicated (n=97). For the treatment-indicated subgroup, this PAI threshold corresponded to a 30% reduction in average post-treatment PANSS scores for patients in the treatment arm relative to the scores of patients in the placebo arm and an improved treatment effect size (Cohen’s d = 1.51 relative to the full sample d = 0.82). Note that this treatment effect size is just an intermediate calculation as PAI is not the only step in our approach given that it does not provide the level of explainability that BRL does. An evaluation of only the PAI step’s out of sample generalization and cross-validated treatment effect improvement is provided in the Supplementary Materials under the PAI Validation section.

**Figure 2:**
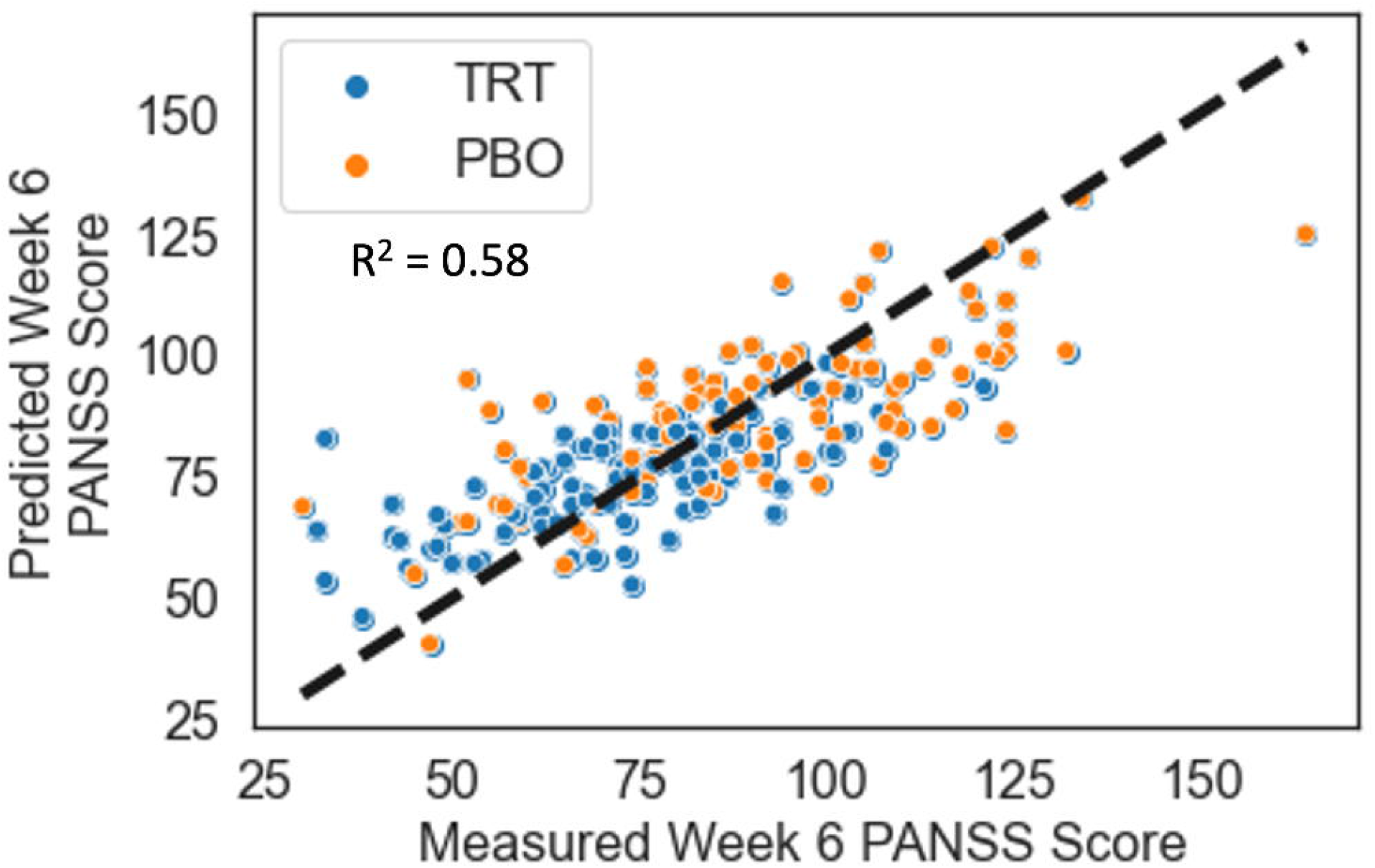
Individual PAI prediction results for actual randomized arms. The plot shows the measured total PANSS score at week 6 vs. the averaged predicted total PANSS score at week 6 from the Elastic Net regression model (each dot is an individual patient, n=197). Patients are colored by their actual randomized arm (paliperidone treatment in blue, placebo in orange), and as expected the week 6 scores are generally higher for patients receiving placebo. The dashed line is y=x. Variance explained by the model is 58%. Note that these are not the counterfactual predictions.

**Figure 3:**
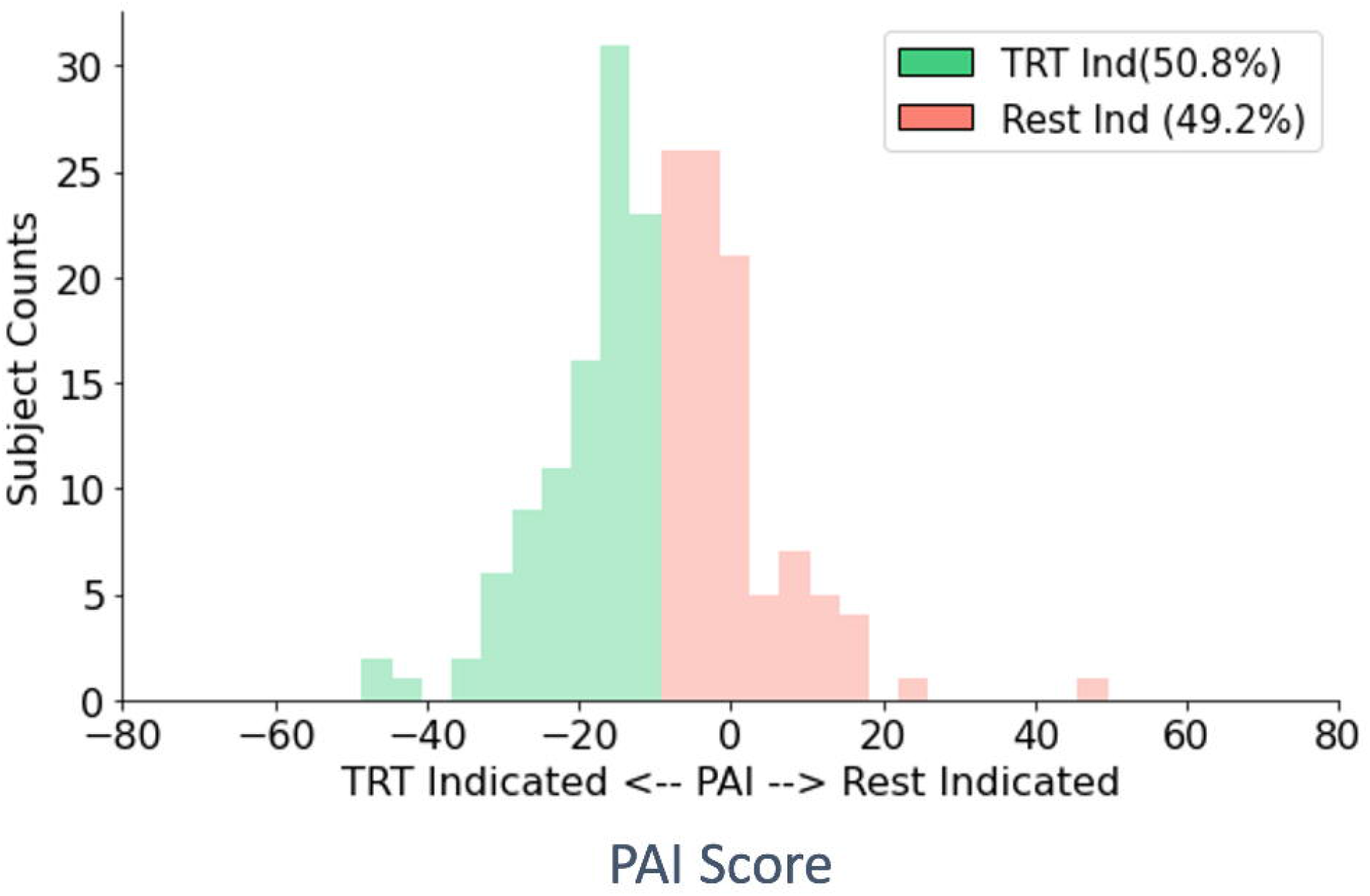
PAI score thresholded graph. A threshold of −9.5 was chosen to create roughly balanced classes and indicates that membership in the treatment-indicated subgroup required a predicted treatment arm PANSS score that is 9.5 points less than the predicted placebo arm PANSS score.

After determining the indication labels from the thresholded PAI scores, we trained the BRL classifier on these labels using 5-fold cross-validation. For the training data, the BRL classifier had an average accuracy of 77.9% (standard deviation = 2.0%), an average Area Under the ROC Curve (AUC) of 0.83 (std = 0.02), and an average F1 score of 0.77 (std = 0.02) across the five folds. For the test data, the BRL classifier had an average accuracy of 74.1% (standard deviation = 5.1%) an average AUC of 0.76 (std = 0.04), and an average F1 score of 0.73 (std = 0.04) across the five folds. Of the 197 patients from the full randomized sample, 87 were labeled treatment-indicated by the BRL algorithm, and the full confusion matrix of cross-validated labels are shown in Figure 4. On the full cross-validated test results, the overall accuracy was 74.1%, the AUC score = 0.74, and the F1 score = 0.73.

**Figure 4:**
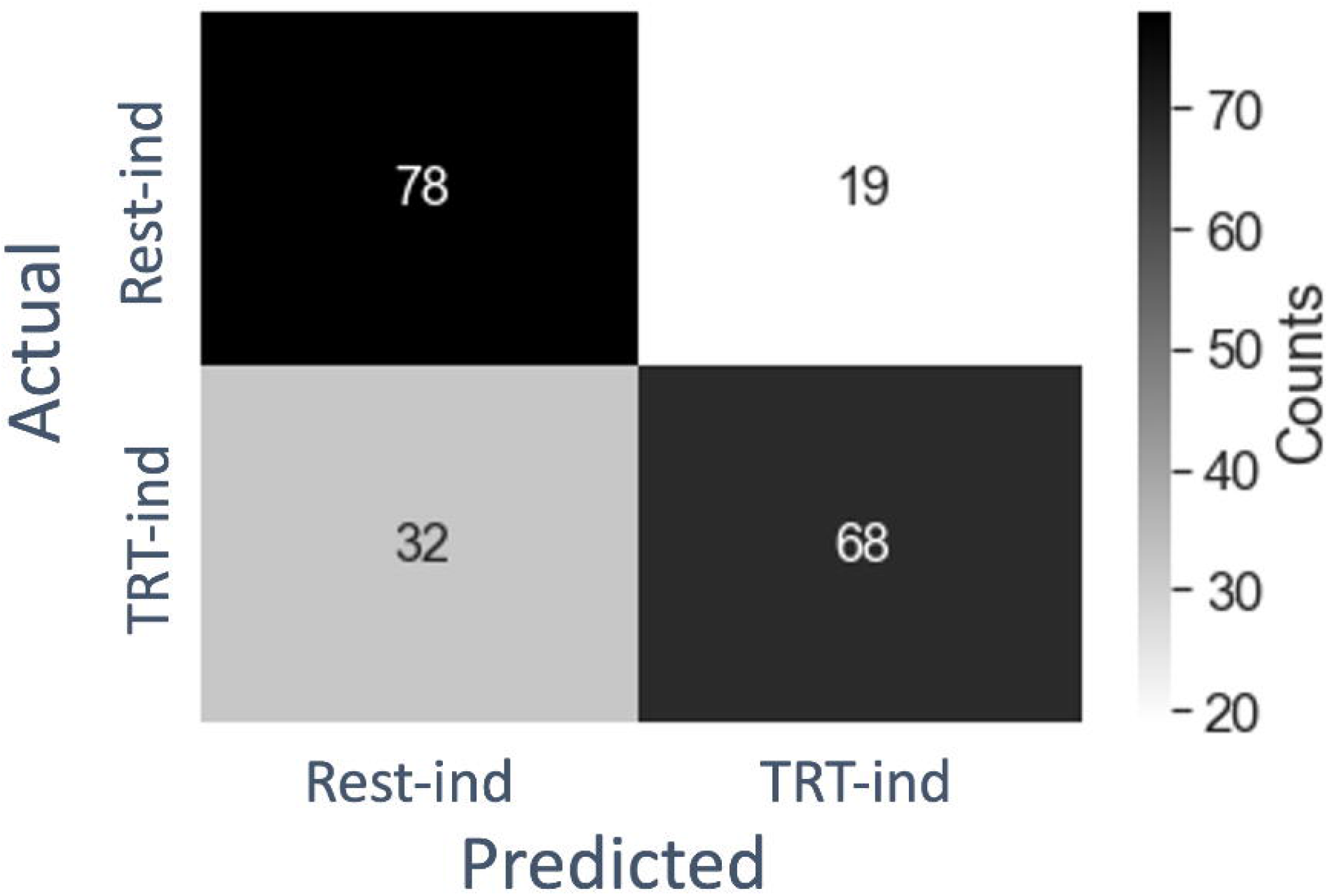
Confusion matrix for cross-validated BRL labels. Actual labels are PAI-derived labels, and predicted labels are BRL-derived labels.

Comparison of the two arms (treatment, placebo) for the full sample v. the treatment-indicated group shows an increase in the Cohen’s d between arms from 0.82 to 1.24 (Figure 5). We also assessed if this large increase in effect size was consistent for the BRL-classified treatment-indicated subgroup and if it was significantly greater than the full sample effect size. We generated a distribution of Cohen’s d’s by performing the same BRL process 100 times and calculating the treatment effect for the treatment-indicated subgroup each time. We found that the treatment effects from these 100 iterations of subgrouping were statistically greater than the full sample treatment effect (BRL-classified treatment-indicated subgroup Cohen’s d’s mean = 1.22, std = 0.09; two-sided, 1-sample t-test: t=43.2, p<0.0001).

**Figure 5:**
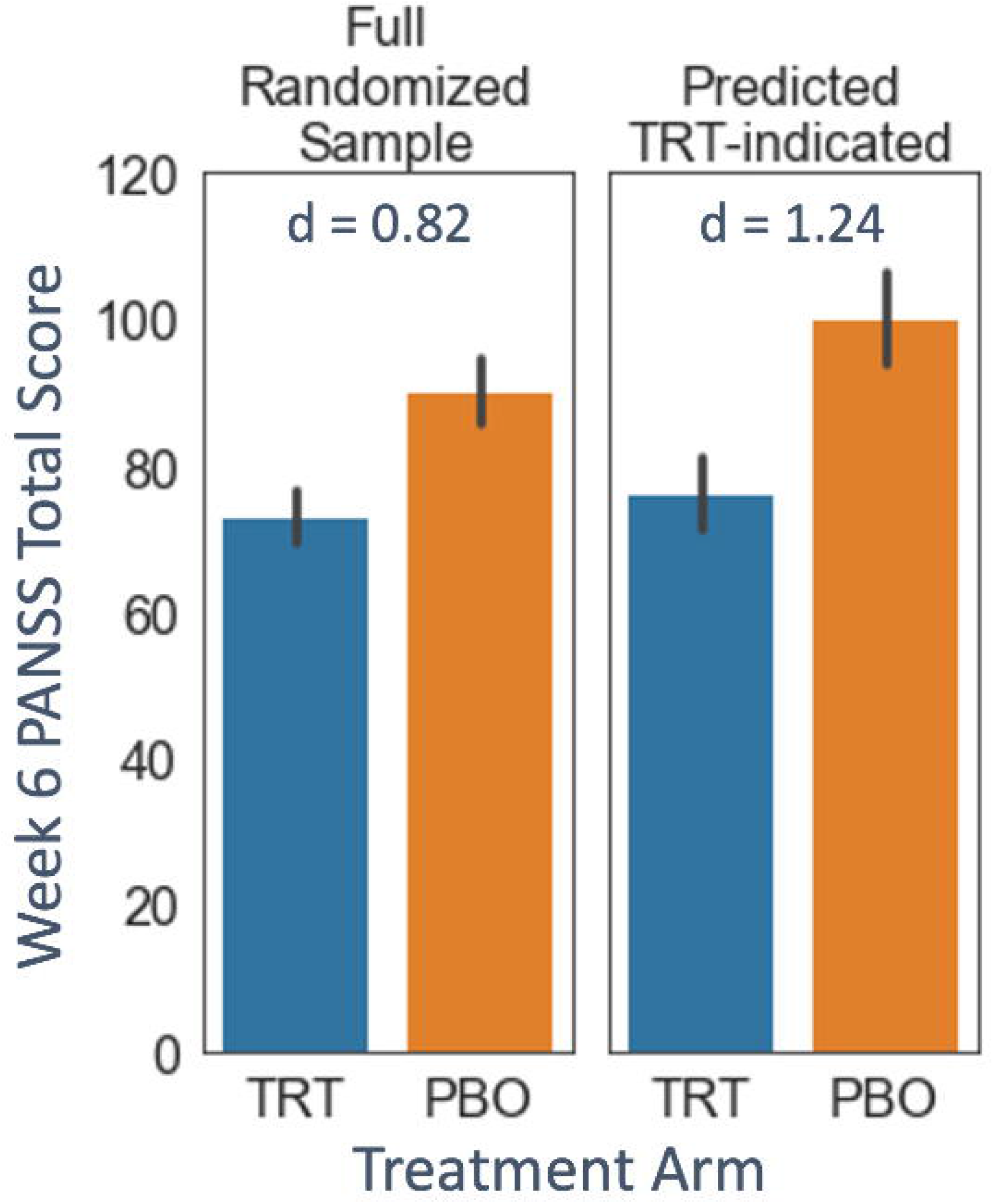
Comparison of actual post-treatment PANSS scores for the full sample and the treatment-indicated subgroup. Bars display treatment (TRT) and placebo (PBO) arms for the full randomized sample (left graph, TRT n=95, PBO n=102) with an illustrative instance of the BRL-classified treatment-indicated (TRT-indicated) subgroup (right graph, TRT n=41, PBO n=46). At a Cohen’s d of 1.24, the effect size between arms for the treatment-indicated subgroup is increased more than 50% over the effect size of the full sample (d = 0.82). Error bars are 95% confidence intervals on the mean.

The five rule lists returned by the BRL classifier differ slightly across the five folds but did identify high disturbance of volition and high uncooperativeness as commonly identifying baseline features of patients who were more likely to respond on treatment than on placebo. Table 2 displays the five rule lists.

**Table 2:**
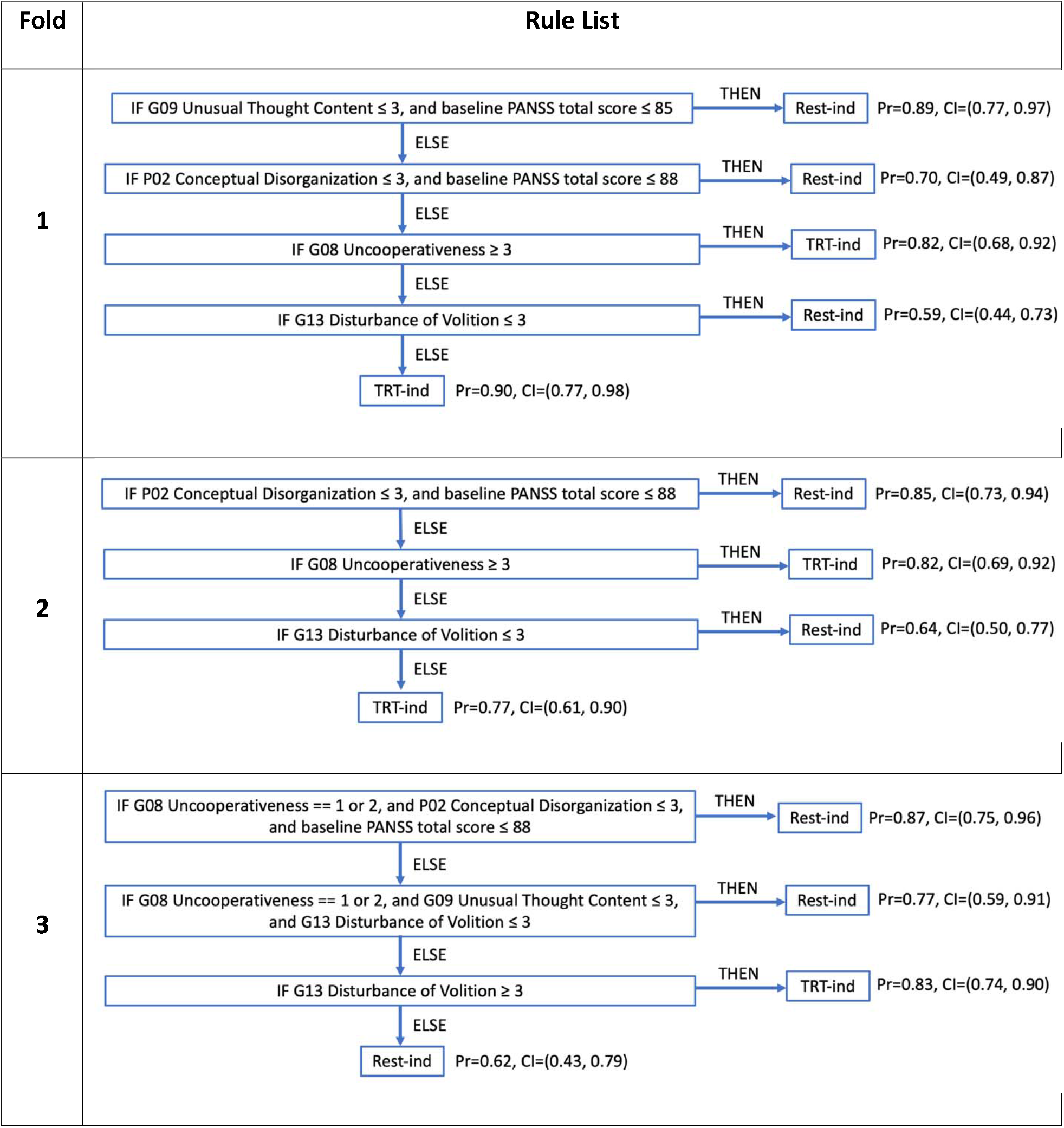
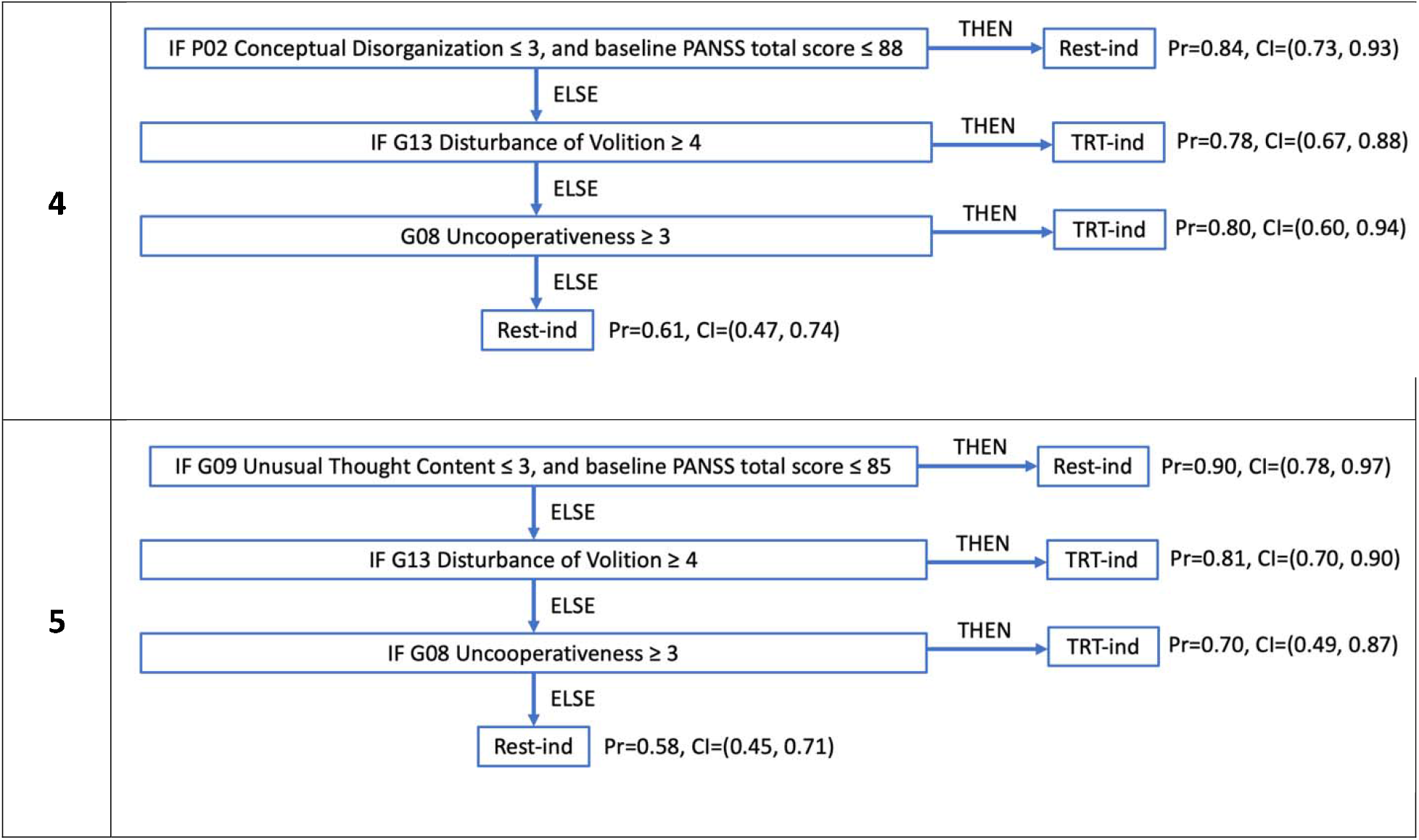
List of the five rule lists created by the BRL model across the five folds. The probability shown in parentheses after each rule is estimated from the percent of patients who satisfy that rule and were labeled with the given label for the rule by the PAI “ground-truth” labels, and the confidence intervals were estimated with bootstrapping. The alphanumeric symbol before each symptom (e.g., P02, G08) refers to the question number from the PANSS scale. Two individual item scores from the PANSS scale repeatedly were involved in subtyping the treatment-indicated patients. Both Disturbance of Volition >= 3 and Uncooperativeness >= 3 appear in multiple rule lists for the treatment-indicated (TRT-ind) subgroup and are categorized as General Psychopathology symptoms.

Though the rule lists show some differences across folds, we found that they still classified patients similarly when applied to the whole data set (not just the test set). To quantitatively assess the consistency of rule list classification, we classified each patient five times with the five rule lists. This gave us five labels (either treatment-indicated or rest-indicated) for each patient from which we can calculate the number of times that a patient is classified as treatment-indicated (max possible is five times). Figure 6 shows a histogram of the number of times that patients were labeled treatment-indicated. For consistent classifiers, most patients should be labeled treatment-indicated either five times or zero times (which corresponds to a patient labeled rest-indicated five times), and the labeling reflects this well according to our histogram. Additionally, most patients labeled by the BRL algorithm treatment-indicated four or five times were treatment-indicated according to the “ground truth” PAI labels. As expected, this was reversed with most patients labeled by the BRL algorithm treatment-indicated zero times were rest-indicated according to the “ground truth” PAI labels. Thus, the five rule lists mostly subtype patients similarly though their wording can differ.

**Figure 6:**
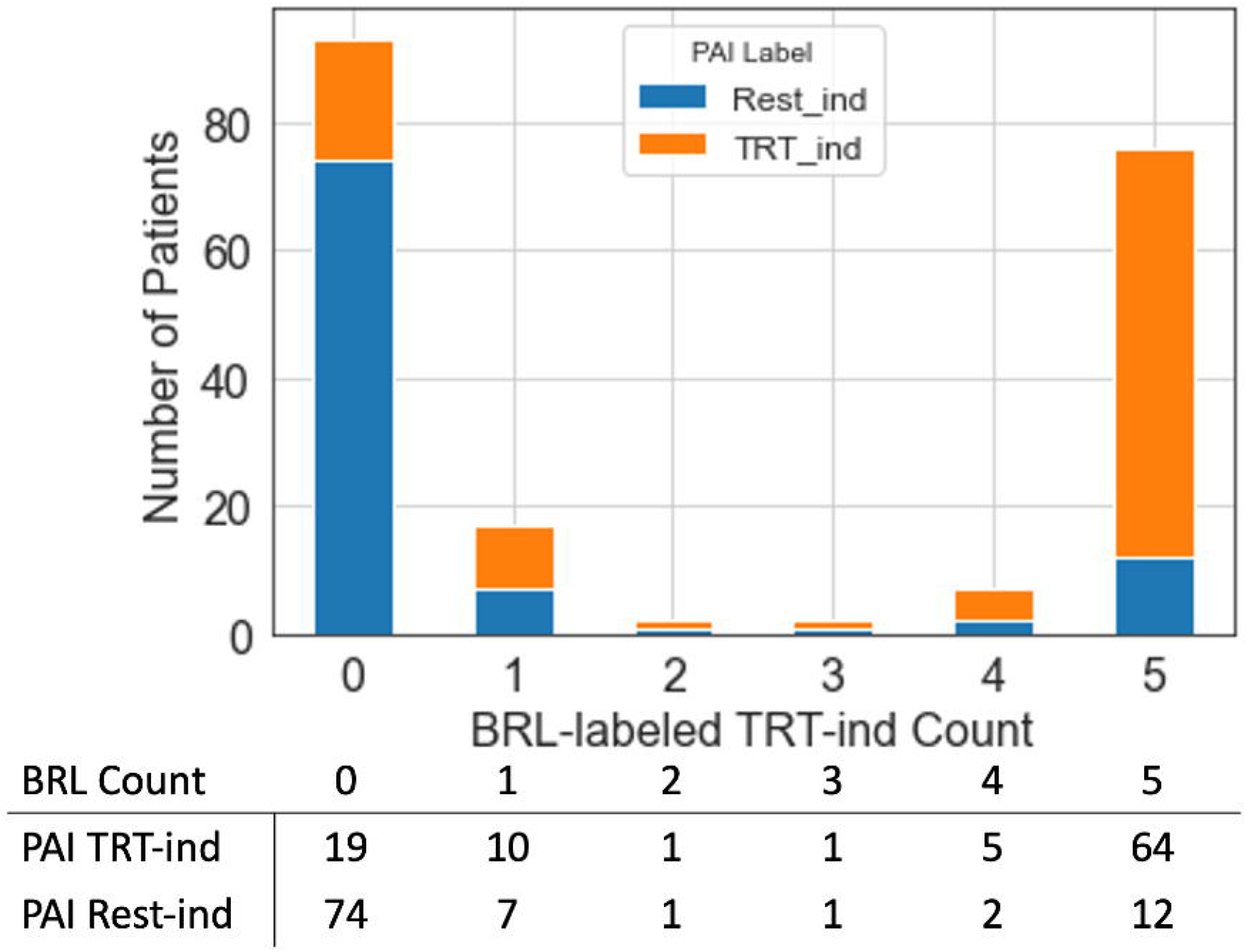
The treatment-indicated (TRT-ind) labeling consistency is seen with a histogram of BRL-labeled treatment-indicated counts. It reflects the number of times (count) that a patient was classified as treatment-indicated across all five rule lists from the 5-fold BRL cross-validation. Most patients are either classified five times or zero times which indicates a higher level of consistency in patient subtyping across the different rule lists. Additionally, most patients in the 5-count column were also labeled as TRT-ind by the PAI algorithm (orange portion of the bar) while most patients in the zero-count column were labeled as Rest-ind by PAI (blue portion of the bar). Patient numbers corresponding to the orange and blue portions are shown in the table below the histogram.

We additionally generated a final BRL model that could be validated on external data sets (Figure 7). This model generated a single rule list as it is trained on the full data set as opposed to the cross-validation approach which generated five rules lists across the five folds. High baseline uncooperativeness and high baseline disturbance of volition each remain predictive of treatment-indicated subgroup membership. For training and testing on the full data set, accuracy was 76%, AUC was 0.81, and the F1 score was 0.75.

**Figure 7:**
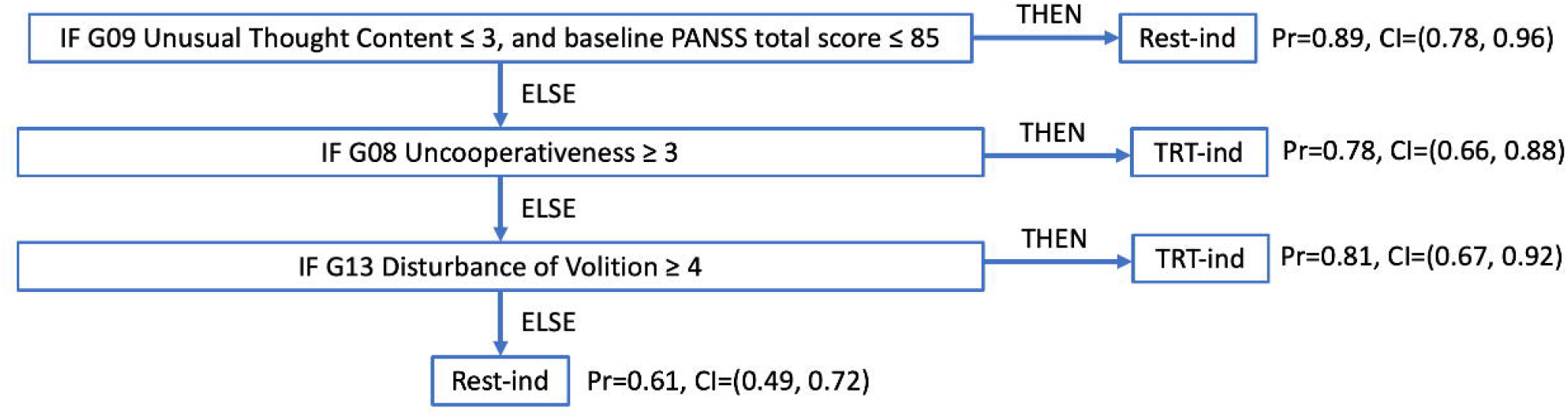
“Final” rule list created by the BRL model when trained on the full data set. The two individual item scores for Disturbance of Volition and Uncooperativeness again appear to classify the treatment-indicated (TRT-ind) subgroup.

## 4. Discussion

With this retrospective analysis, we have technically-validated an approach of using a combination of machine learning methods to identify clinically-explainable rules that effectively subtype paliperidone-indicated schizophrenia patients who show an improved treatment effect over the full randomized sample. This extends the prior validation that demonstrated the method’s effectiveness in a clinical trial of a novel depression treatment [8].

While this validation was performed on a successful trial where the full sample already displayed the success of the experimental drug, we demonstrated than the treatment effect can be further improved with a patient selection approach. Statistically speaking, increasing the effect size can help decrease the enrollment numbers for patients, thereby possibly decreasing the cost and time of a new clinical trial (for example when running a more targeted confirmatory target phase two trial after gathering data in a traditional phase two trial). Thus, in addition to patient selection that improves the treatment effect for unsuccessful trials [5, 8], our results suggest that patient selection could help clinical development even for treatments with stronger effects.

While the primary goal in this study was to validate a patient selection approach, the methodology also allows us to better understand a potential paliperidone-responsive subtype of schizophrenia. A majority of the rule lists generated from the cross-validation framework found two general psychopathology symptoms, disturbance of volition and uncooperativeness, to be predictive of membership in the paliperidone-indicated subgroup suggesting that paliperidone may be indicated for the treatment of patients with either of these symptoms. While subtyping schizophrenia is an active area of research [e.g., 2, 9], the higher severity of uncooperativeness and disturbance of volition seen in this paliperidone-indicated subgroup has not been previously described and should be externally validated. For this reason, we have included a single BRL rule list as a “final” model which could be tested by others interested in a paliperidone-indicated subgroup.

This approach is particularly useful in the context of selecting patients for clinical trial enrollment as the clinical trial outcome is dependent on large effects that are seen for patients in the treatment arm but not placebo arm. However, the proposed approach with the additional clinician-friendly explainability of BRL could make it more broadly useable as a clinical decision support system which are not commonly incorporating machine learning yet [10, 11]. The framework could be extended to accommodate multiple classes for indications of multiple treatments. With the proper validation, this could provide clinicians with a tool to match the best of several possible treatments to a particular patient.

Some limitations with this approach remain. The PAI model does have some bias in its predictions as shown by the greater differences in measured and predicted week 6 PANSS scores in the larger and smaller ranges. This reflects a model that is underfit and could be due to missing predictor variables or due to using a linear rather than non-linear model. Future iterations of this approach could test using a non-linear approach such as generalized random forest [12] to improve predictions in this first step. As the shortcomings of the PAI step can affect model accuracy of the BRL step, there is additional incentive to improve predictions from the PAI step. Even with this weakness, the PAI model still provided adequate predictions to allow the BRL model to find a treatment-indicated subgroup with improved treatment effect size. Another limitation is the testing only within a single data set. A more robust approach would be to test the BRL model in a separate data set to assess generalization of the model and the proposed paliperidone-indicated schizophrenia phenotype. Some may question using the baseline total PANSS score as a predictor variable either due to its possible collinearity with other PANSS item scores or that the resulting use of lower PANSS scores to classify rest-indicated patients (therefore higher PANSS score is indirectly predictive of treatment-indicated patients) may be reflecting an effect of regression to the mean for the treatment-indicated patients. Unpublished analyses in our lab did not find major differences in performance or predictive features whether including or not including this variable. Additionally, the critical result is not that PANSS scores are reduced for the treatment-indicated patients, but that they are reduced much more on treatment than on placebo. Thus selecting patients as rest-indicated based on the baseline total PANSS score and the implications that has for selecting treatment-indicated patients does not have any bearing on the improved difference seen between arms of the treatment-indicated patients. And finally, this modeling approach cannot currently handle longitudinal independent or dependent variables, but extending the framework of this method to more data types could expand its useability.

## 5. Conclusions

These results help to technically validate our explainable AI approach to patient selection for a clinical trial by identifying a subgroup of schizophrenia with an improved treatment effect. Importantly, this approach opens up the possibility of identifying a targeted subgroup of patient prior to randomization to an arm and improving the clinical trial outcome by using this patient subgroup in particular.

## Data Availability

The data that support the findings of this study are available from the YODA Project (https://yoda.yale.edu/) but restrictions apply to the availability of these data, which were used under license for the current study, and so are not publicly available.

## Abbreviations

AI: Artificial Intelligence
AUC: Area Under the ROC Curve
BRL: Bayesian Rule List
FDA: U.S. Food and Drug Administration
PANSS: Positive and Negative Syndrome Scale
PAI: Personalized Advantage Index
PBO: Placebo
ROC: Receiver Operating Characteristic
Rest-ind: Rest-indicated
TRT: Treatment
TRT-ind: treatment-indicated
XAI: Explainable Artificial Intelligence
YODA: Yale University Open Data Access

## Declarations

### Ethics approval and consent to participate

This study makes use of de-identified data made available via the YODA Project (https://yoda.yale.edu; research proposal number 2019-4080) and was exempt from ethical oversight.

### Consent for publication

Not applicable.

### Authors’ contributions

JT, MK, MM, and TL helped conceive of the approach to PAI+BRL validation with the YODA data set and subsequent analysis, and JT advised on methodology with the team who developed the PAI+BRL approach. MM performed the analysis of the data. MM, MK, and TL interpreted the results, and wrote or revised the manuscript. All authors read and approved the final manuscript.

## Acknowledgments

We would like to thank Clark Gao, Yuelu Liu, Humberto Gonzalez, and Parvez Ahammad for their contributions in creating the PAI+BRL methodology that is used here in a different application, and Kathy Tracy, Rezi Zawadzki, Andrew Jaffe, John Dunlop, and Bill Martin for further discussions. This study, carried out under YODA Project # 2019-4080, used data obtained from the Yale University Open Data Access Project, which has an agreement with JANSSEN RESEARCH & DEVELOPMENT, L.L.C. The interpretation and reporting of research using this data are solely the responsibility of the authors and does not necessarily represent the official views of the Yale University Open Data Access Project or JANSSEN RESEARCH & DEVELOPMENT, L.L.C.

