## Supplementary Material for "Explainable AI enables clinical trial patient selection to retrospectively improve treatment effects in schizophrenia"

*Supplementary Materials*

PAI Validation

While PAI is only the first step in our two-step approach to patient selection, we evaluated it as a single step approach to patient selection as a follow up to the PAI validations shown previously ([4], [5], main references). Thus, we wanted to calculate the treatment-indicated subgroup’s treatment effect size that we might expect in unseen samples when using PAI alone.

To evaluate generalization performance to unseen samples, we used a 5-fold cross-validation (CV) framework that generated a regression model on the 80% of training data and used it to predict actual and counterfactual post-treatment scores for the patients in the remaining 20% of test data. The actual score predictions were then used to calculate R^2^ below. Specifically, we used a nested cross-validation approach where an inner loop performs a grid-search (across the range of *alpha* = [0.001, 0.01, 0.1, 1, 10] and *l1_ratio* = [0.1, 0.5, 0.9]) to optimize hyperparameters for the elastic net regression and an outer loop evaluates generalization error. Each time the 5-fold CV is run, the patient subsets are different across the different folds. This returns different results for each run of 5-fold CV PAI (but not the full data set training as done in the main workflow presented in the main manuscript). Thus, we performed this modeling 100 times to assess the average performance. We maintained the PAI score thresholding at the point that labeled 50% of the sample as treatment-indicated as this is an evaluation of the PAI process as outlined in the manuscript’s main workflow (see Figure 1 in the main text). If PAI alone is being used, however, the researcher could prioritize using a clinical cutoff (like 30% improvement of the treatment arm over the placebo arm for the treatment-indicated subgroup) and adjust the threshold accordingly to improve the treatment effect.

We found statistically significant differences in both the R^2^ for predicted post-treatment scores of the actual arm from the regression modeling (mean R^2^ =0.25, std R^2^ =0.02; two-sided, 1-sample t-test where the null hypothesis is that R^2^ = 0: t = 118.28, p<0.0001) and the Cohen’s d of the treatment-indicated subgroup after PAI thresholding (mean d = 0.96, std d = 0.14; two-sided, 1-sample t-test where the null hypothesis is that d = 0.82 from the full randomized sample treatment effect: t = 10.17, p<0.0001). These out-of-sample results generated with a cross-validation framework suggest the possibility of PAI generalization on additional external data, but this should be validated explicitly with an external data set.
